# Longitudinal change in mitochondrial heteroplasmy exhibits positive selection for deleterious variants

**DOI:** 10.1101/2024.03.27.24304930

**Authors:** Lieke M. Kuiper, Wen Shi, Joost Verlouw, Yun Soo Hong, Pascal Arp, Daniela Puiu, Linda Broer, Jiaqi Xie, Charles Newcomb, Stephen S. Rich, Kent D. Taylor, Jerome I. Rotter, Joel S. Bader, Eliseo Guallar, Joyce B.J. van Meurs, Dan E. Arking

**Affiliations:** Genetic Laboratory, Department of Internal Medicine, Erasmus MC, Rotterdam, the Netherlands; McKusick-Nathans Institute, Department of Genetic Medicine, Johns Hopkins University School of Medicine, Baltimore, MD, USA; Department of Biomedical Engineering, Johns Hopkins University, Baltimore, MD, USA; Center for Public Health Genomics, Department of Public Health Sciences, University of Virginia, Charlottesville, VA, USA; The Institute for Translational Genomics and Population Sciences, Department of Pediatrics, The Lundquist Institute for Biomedical Innovation at Harbor-UCLA Medical Center, Torrance, CA, USA; Department of Epidemiology and Medicine, and Welch Center for Prevention, Epidemiology, and Clinical Research, Johns Hopkins University Bloomberg School of Public Health. Baltimore, MD, USA; Department of Orthopeadics & Sports Medicine, Erasmus MC, Rotterdam, the Netherlands

## Abstract

A common feature of human aging is the acquisition of somatic mutations, and mitochondria are particularly prone to mutation due to their inefficient DNA repair and close proximity to reactive oxygen species, leading to a state of mitochondrial DNA heteroplasmy^1,2^. Cross-sectional studies have demonstrated that detection of heteroplasmy increases with participant age^3^, a phenomenon that has been attributed to genetic drift^4–7^. In this first large-scale longitudinal study, we measured heteroplasmy in two prospective cohorts (combined n=1405) at two timepoints (mean time between visits, 8.6 years), demonstrating that deleterious heteroplasmies were more likely to increase in variant allele fraction (VAF). We further demonstrated that increase in VAF was associated with increased risk of overall mortality. These results challenge the claim that somatic mtDNA mutations arise mainly due to genetic drift, instead demonstrating positive selection for predicted deleterious mutations at the cellular level, despite an negative impact on overall mortality.

---

Mitochondria are involved in essential cellular processes, such as ATP and iron-sulfur cluster synthesis, cell calcium homeostasis, and apoptosis^1^. They possess their own unique genome, with each cell containing hundreds to thousands of mitochondrial DNA (mtDNA) copies^1,2^, and due to limited repair capacity, mtDNA is highly mutable, with an estimated five-to-fifteen-fold higher mutation rate than nuclear DNA^8–10^. The elevated mutation rate contributes to the high levels of inherited mtDNA diversity and the generation of somatic mtDNA mutations. Inherited mutations in mtDNA can lead to mitochondrial dysfunction and they have been associated with aging-related phenotypes such as longevity^11^, cancer^12^, and neurodegenerative diseases^13^. Somatic mtDNA mutations, acquired throughout an individual’s lifetime^1^, often result in mitochondrial heteroplasmy, a state of mutations being present in a subset of mtDNA molecules within a cell or tissue. These mutations are generally believed to be a product of random genetic drift^4–7^, however, the presence of mtDNA heteroplasmy, measured cross-sectionally, has been associated with all-cause mortality and cancer in ∼200,000 participants in the UK Biobank^3^, raising the question of whether somatic heteroplasmies are under selection. Indeed, in the absence of longitudinal studies, our understanding of the dynamics of mtDNA heteroplasmy in the general population remains limited. We therefore assessed mitochondrial heteroplasmy in two large population-based studies across time, with two primary objectives: 1) to determine whether somatic heteroplasmy arises as a function of genetic drift or natural selection; and 2) to determine whether change in heteroplasmic variant allele fraction (VAF) is associated with health outcomes.

To address these questions, we measured mtDNA heteroplasmy at two time points in DNA derived from buffy coat in participants from two population-based prospective cohorts: the Multi-Ethnic Study of Atherosclerosis (MESA) (n=1031; median baseline age = 58.5 years; median time between measurements of 9.4 years)^14^ and the Rotterdam Study (RS) (*n*=373; median baseline age = 64.8 years; median time between measurements of 6.4 years)^15^. Study population characteristics are shown in **Supplementary Table 1**. We set a heteroplasmy variant allele frequency (VAF) threshold of 5-95%, chosen a priori to maximize sensitivity to true heteroplasmies, while minimizing false-positives due to nuclear-encoded mitochondrial sequences (NUMTs)^3^. We classified a heteroplasmy as de novo or lost only when the heteroplasmy was below the detection limit (VAF= 5%) in the first or second visit, respectively, and above the detection limit in the other visit. This criterion held even in cases of variants with low read depth, ensuring a reliable assessment of heteroplasmy changes between visits. The main analysis focused on calculating deltaVAF measured as the change in rate of growth between the heteroplasmy and reference allele between visits: 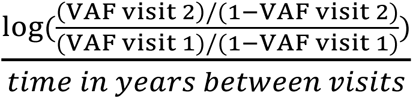. If the absolute value of deltaVAF was <0.05, we considered the heteroplasmy to be stable.

In MESA, we observed 388 participants with heteroplasmies at either visit; 331 (32.1%) and 358 (34.7%) at the initial and follow-up visits, respectively. Along with the increase in heteroplasmic individuals, we also observed an increase in participants harboring >1 heteroplasmy at the follow-up visit (81 vs. 58 in 1st visit). In RS, we observed 152 participants with heteroplasmies at either visit; 129 (34.6%) and 142 (38.1%) at the initial and follow-up visits, respectively. In RS, we observed a small increase in the number of participants with >1 heteroplasmy in the second visit (31 vs 33). While VAF of heteroplasmic variants was highly correlated across visits (MESA Pearson’s r^2^=0.80, RS Pearson’s r^2^=0.79) (**Figure 1a,b**), there was a notable upward shift in VAF in the later visits (**Figure 1c,d**). Of the 110 de novo heteroplasmies observed in MESA and 39 in RS, none was completely de novo (i.e., no alternate read at prior visit), indicating that the change in heteroplasmy count is entirely explained by a change in VAF among mutations that already existed prior to the initial visit.

**Fig 1.**
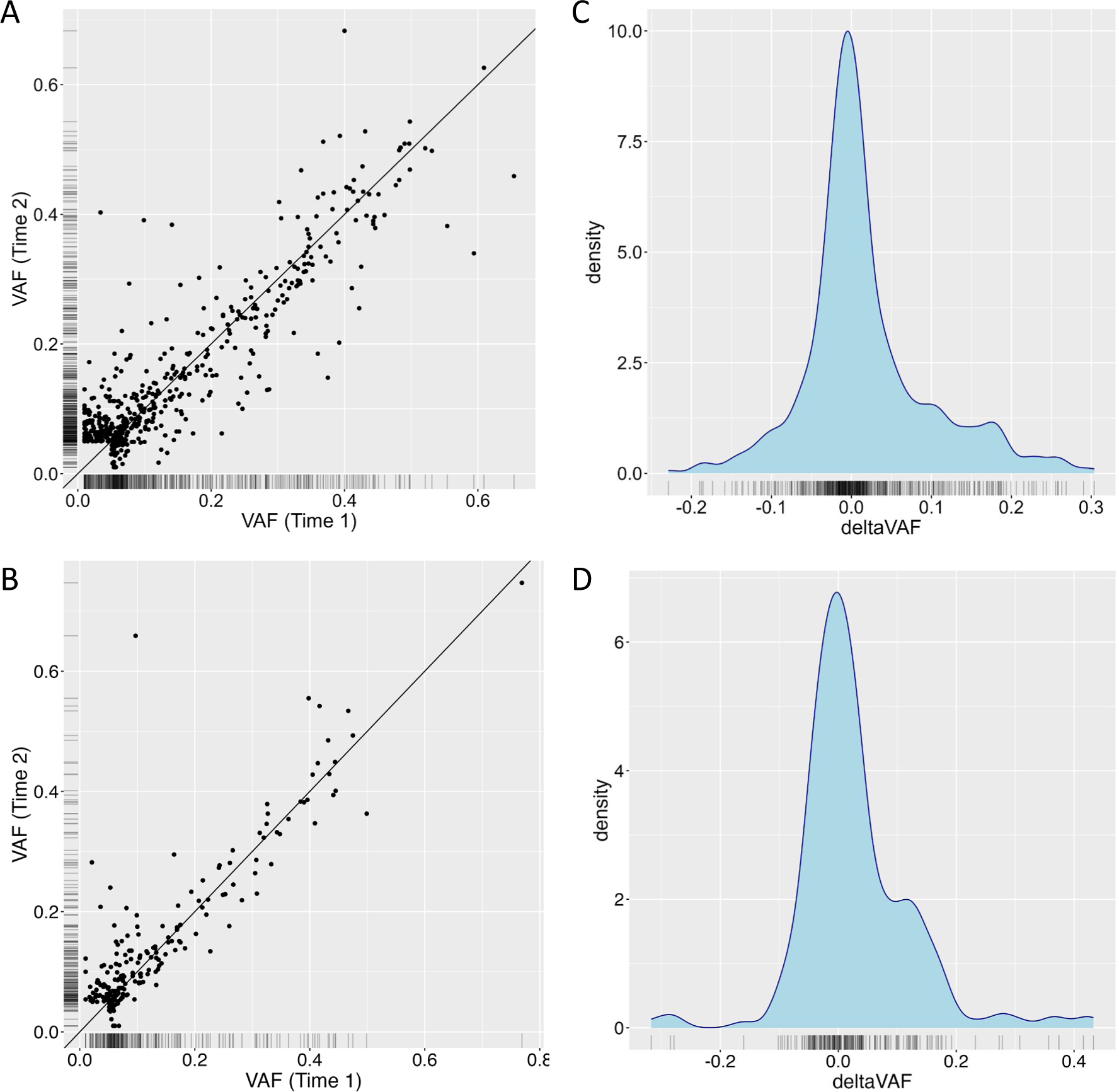
Change in variant allele fraction (VAF) over time. **a,b**, Correlation between VAF at the first measurement (x-axis) and the second measurement (y-axis), with the black line indicating perfect correlation, in MESA (**top**) and RS (**bottom**). **c,d**, Distirbution of the change in VAF (deltaVAF) in MESA (**top**) and RS (**bottom**).

To determine whether natural selection was playing a role in the increased VAF observed with aging, we first stratified heteroplasmies by functional categories (**Figure 2a,b**). Compared to synonymous and D-loop mutations, in MESA we observed a significant increase in deltaVAF in missense mutations (*P*<0.003) and RNA (*P*<0.0005) heteroplasmies (**Supplementary Figure 1a**). Despite a smaller sample size, consistent effect estimates were observed in RS for missense heteroplasmies (*P*<0.032), and a meta-analysis across both cohorts showed significantly higher deltaVAF for both missense (*P*<0.0002) and RNA (*P*<0.001) heteroplasmies relative to synonymous and D-loop variants, suggesting non-random expansion of mitochondrial heteroplasmies (**Supplementary Figure 1a**). To further test whether functional heteroplasmies were increasing in VAF, we made use of a modified version of a recently developed score (mMLC score, see **Online Methods**) that uses local constraint to estimate the functional consequences of mitochondrial heteroplasmies. The mMLC ranges from 0 to 1, with 0 indicating a benign mutation and 1 corresponding to a nonsense mutation that completely abrogates protein function^16^. In both MESA (*P*<5.0×10^−7^) and RS (*P*<0.035), we saw a significantly higher deltaVAF associated with higher mMLC score (**Figure 2 c,d**), indicating that predicted deleterious variants are more likely to increase in VAF with aging (meta-analysis P<5.0×10-8, **Supplementary Figure 1b**). To address potential confounding due to non-driver (i.e., passenger) mutations, we also limited analyses to participants with only a single heteroplasmy, or by choosing a single heteroplasmy per participant based on the highest MLC score, and obtained similar results (**Supplementary Figure 2**).

**Fig 2.**
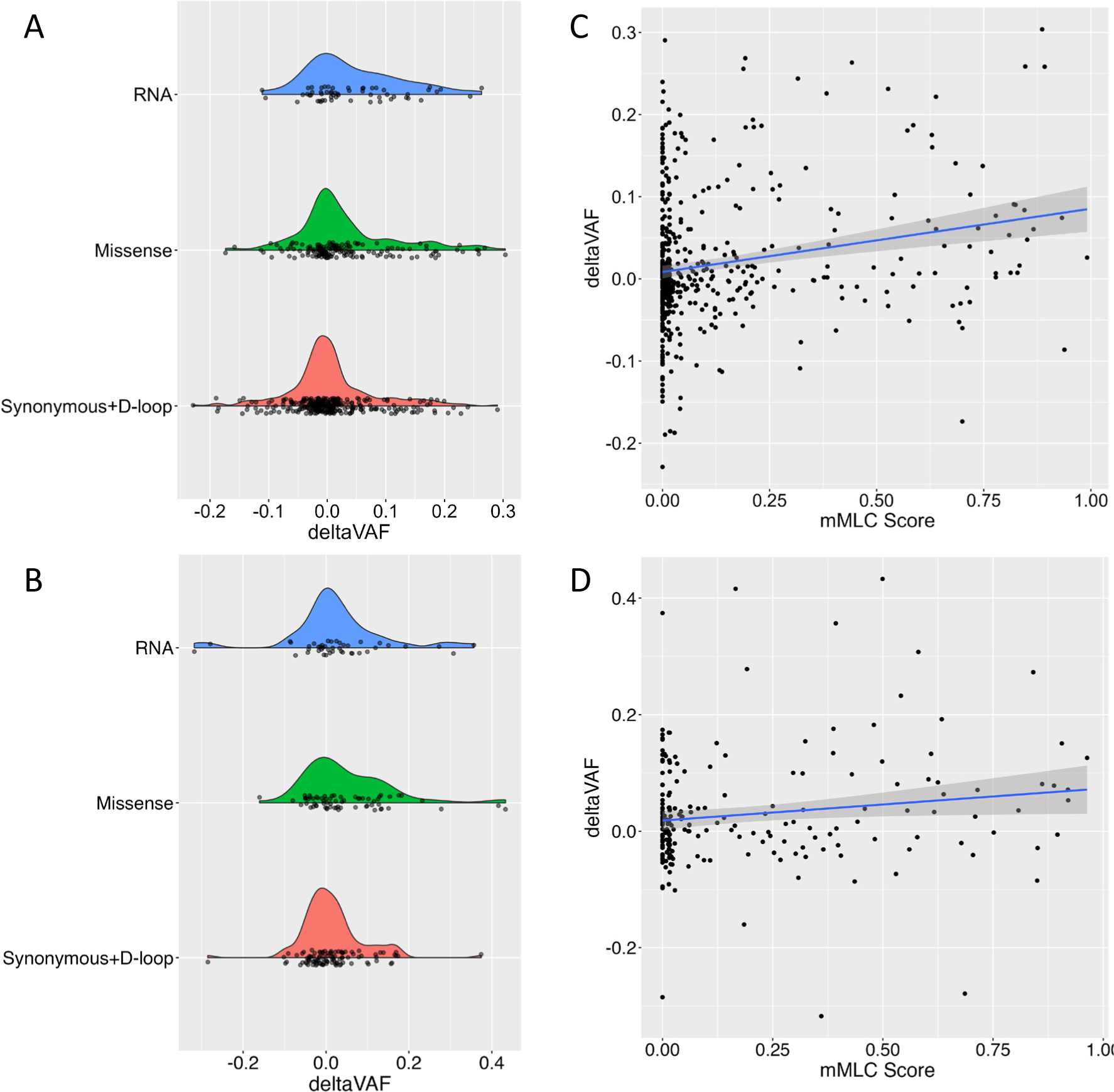
deltaVAF by functional consequence. **a,b**, Density plots of deltaVAF stratified by mutation consequences in MESA (**top**) and RS (**bottom**). **c,d**, Correlation between the mMLC score and deltaVAF, with a blue line with confidence intervals illustrating the regression slope in MESA (**top**) and RS (**bottom**).

Previous work has shown that mitochondrial heteroplasmy burden, measured as the MLC score sum (MSS) across all heteroplasmies in a given individual, is associated with an increased risk of overall mortality^3^. To determine if the change in VAF is likewise associated with overall mortality, for each individual we selected the variant with the largest increase in deltaVAF, and assessed the association with overall mortality (**Figure 3**). In MESA, there were 53 deaths out of 385 samples with follow-up mortality data, with a median follow-up time of 8.1 years (Q1=7.8, Q3=8.4). After adjusting for age, sex, collection center, race, and smoking status, a 0.10 increase in deltaVAF was associated with a 1.52-fold (95% CI 1.09-2.11, P<0.014) increased risk of overall mortality. Additionally adjusting for mMSS calculated at the follow-up visit only modestly attenuated the effect (HR=1.42, 95% CI 0.98-2.06). In the RS, there were 51 events among 152 individuals followed for up to 10 years. Highly concordant effect sizes were observed in RS (HR=1.38, 95% CI 1.04-1.82, *P*<0.027), again with only modest attenuation when including MSS into the model (HR=1.29, 95% CI 0.95-1.75). A meta-analysis demonstrates a highly significant association between deltaVAF and mortality (HR=1.43, 95% CI 1.16-1.78, *P*<0.001).

**Fig 3.**
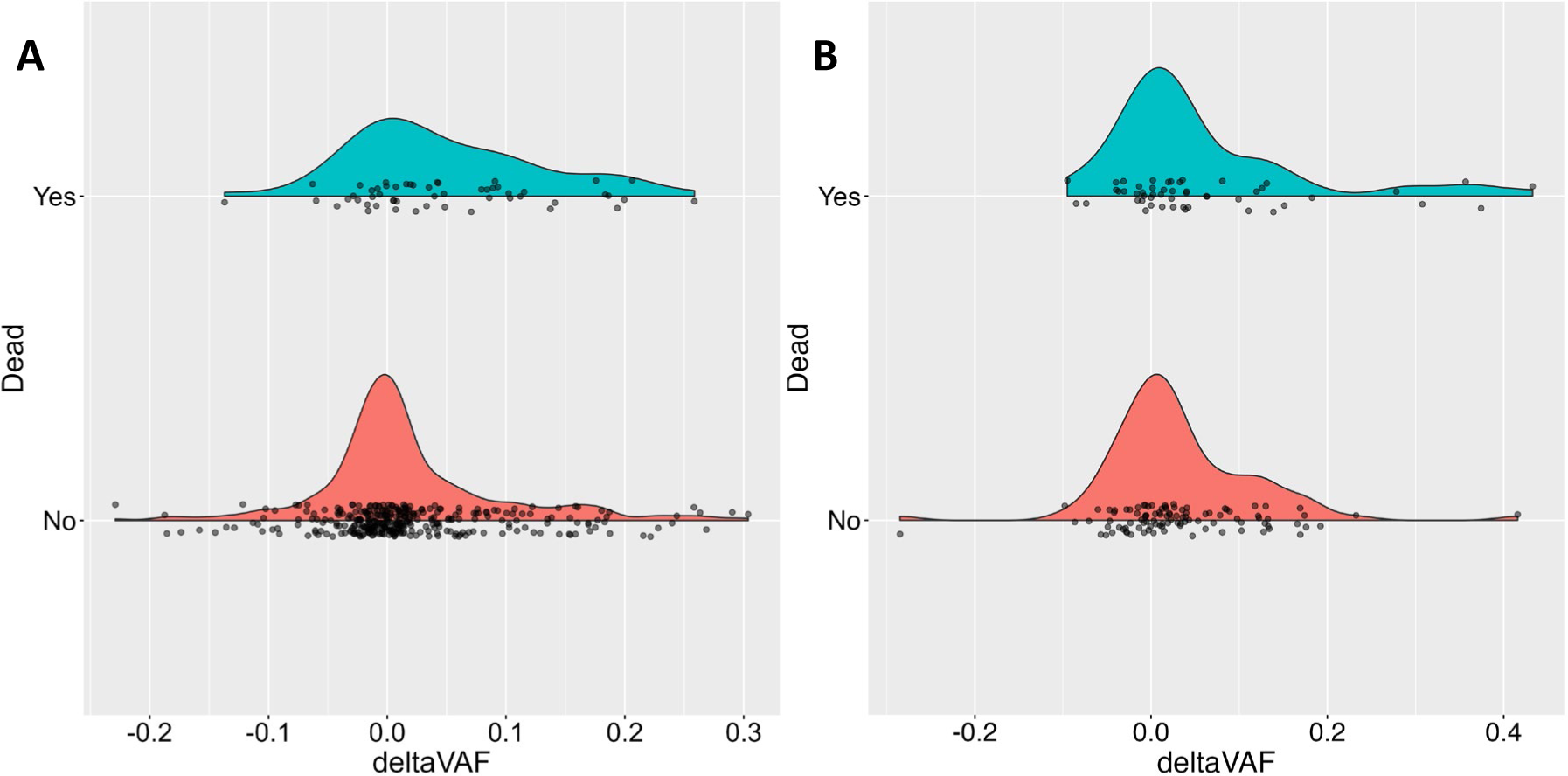
deltaVAF by vital status. Density plots of deltaVAF by vital status in (**a**) MESA and (**b**) RS.

In this first large-scale study of longitudinal mtDNA heteroplasmy measurements in humans from two prospective cohorts, we observed that heteroplasmies are largely stable over time (<0.05 change in deltaVAF, MESA = 325/504 [64.4%]; RS = 131/211 [62.1%]). However, overall, there is an increase in VAF, largely driven by variants below detection (5% VAF) at the initial visit (MESA = 90/119 [75.6%], RS=32/59 [54.2%]) (**Supplementary Figure 3**,**Supplementary Table 2**). Variants with large deltaVAF are more likely to be deleterious and are associated with overall mortality. To confirm that these results are not confounded by de novo mutations in our dataset having different mMLC score distributions than extant variants, we re-called heteroplasmies with a VAF cut-off of 2%, and excluded de novo variants (i.e., we only examined heteorplasmies that were detectable at baseline). In both MESA (P<2.30×10^−14^) and RS (P<2.79×10^−4^), higher mMLC score of extant heteroplasmies was associated with greater increase in VAF. These results suggest that natural selection, where heteroplasmies that are deleterious for the organism (human) nevertheless have a survival advantage leading to clonal expansion, plays a significant role in mitochondrial heteroplasmy, challenging the prevailing notion that acquired mitochondrial heteroplasmy arises mainly due to random genetic drift.

Strengths of the current study include the robust results in two large prospective population-based cohorts with longitudinal measures of mitochondrial heteroplasmy. Further, MESA included a majority of non-European ancestry individuals (∼59%), while RS was almost entirely European ancestry, suggesting generalizability of results. As limitations, we note that we only measured mtDNA heteroplasmy at two time points, not allowing for the observation of any non-linear changes in VAF, and the ∼3 year shorter follow-up time in RS that likely contributed to the proportionally fewer de novo variants detected than in MESA (18% vs 22%) (**Supplementary Table 2**).

## Online Methods

### Study populations

#### Multi-Ethnic Study of Atherosclerosis (MESA)

MESA is a prospective cohort of 6,452 individuals aged 45-84 who were free of clinical cardiovascular disease at time of recruitment from six communities across the United States. Participants had examinations at field centers with initial examination in 2000-2002 and exam 5 in 2010-2011^17^. Information collected at exams includes age, sex, race, weight, height, blood pressure, smoking status, medication, lifestyle, family history, and medical history. Blood is drawn from participants at each exam and DNA is extracted from the buffy coat of the first examination and the fifth examination (N=4475). Annual follow-up were conducted through 2019 with mail, electronic mail, and telephone interviews; classification of cardiovascular events and mortality were collected from death certificates, hospital records, and interviews with physicians, relatives, or friends^17^. For the current study, 1105 participants were selected randomly from participants with DNA available from visits 1 and 5 and who did not have a cardiovascular event prior to visit 5.

#### Rotterdam Study

The Rotterdam Study is a population-based cohort with four sub-cohorts located in Ommoord, a suburb of Rotterdam, the Netherlands^15^. The participants of the first sub-cohort (RS-I) consisted of individuals aged ≥ 55 years (n = 7983, response rate 78%) and they visited the research center for the first time between 1990 and 1993. In 2000–2001, individuals who had reached 55 years of age or moved into the study area (n = 3011, response rate 67%) were invited to join the study and the second sub-cohort RS-II was initiated. Between 2006-2008, a third sub-cohort (RS-III) was established enrolling new individuals aged ≥ 45 years (n = 3932, response rate 65%). The fourth cohort started between 2016-2018, consisting of all residents of Ommoord aged 40 years and over who had not been invited previously (n=3005, response rate 45%). Participant information was collected every 3–5 years through home interviews, questionnaires, and examinations at our dedicated research center located in the Ommoord district. The home interviews collected information such as sex, smoking status, and medical history. Blood was drawn when participants visited the research center. Information on the vital status of participants was obtained biweekly via municipal population registries and databases of general practitioners and hospitals^15^. We determined genetic ancestry based on information from all participants of the first RS cohort with available genetic information. The cleaned genotype data from all samples from RS-I was merged with HapMap CEU release 22 (build 36)^18^ (to provide a backbone population set). The genotype data was pruned to only keep variants in linkage equilibrium and then analyzed using *ADMIXTURE* (default parameters used)^19^. We performed cross-validation for 1 to 8 ancestral populations recognized in HapMap. Finally, ancestral groups were delineated based on containing at least 50% genetic material from a specific ancestral group. In participants where genetic ancestry information was unavailable, we assigned them to an ancestry if at least 3 of their grandparents were born in a region. In the current study, 373 participants from the first (RS-I-1) and third (RS-I-3) visit of the first subcohort of the Rotterdam Study with information on heteroplasmy from two time points were included.

### WGS data

For MESA, WGS data from Trans-Omics for Precision Medicine (TOPMed) Freeze 9 data release was available for all first visit samples. The study provides ∼30X coverage WGS data sequenced using Illumina’s next-generation sequencing technology and passed all quality control metrics^20^.

### mtDNA sequencing

#### Rotterdam Study

The Twist Library Preparation EF 2.0 protocol was performed on a Perkin Elmer Sciclone robot with 100 ng DNA and 7 PCR cycles. In total, 1536 different indexes were used. After the library preparation, the DNA concentrations were measured with nanodrop and 16 samples with 200 ng DNA of each sample were pooled for the hybridization step. The pooled DNA was dried using vacuum pressure. To each dried sample, 4 ul of the mtDNA probe panel (Twist) was added and hybridized in a PCR machine at 70C for 17 hours. The capture of the mtDNA was performed on a Perkin Elmer robot with the Twist Target enrichment standard hybridization v2 protocol. For each pool of samples, 15 PCR cycles were performed. DNA concentrations were measured with a fluorescence-based assay and size distribution was checked with the Tapestation system. Sequencing was performed at 2 × 150 bp on an Illumina Novaseq6000 system at up to 10,000x per sample.

#### MESA

The Twist Library Preparation EF 2.0 protocol was performed manually on 50 ng DNA per sample with enzymatic fragmentation at 37C for 20 minutes. Samples were amplified for 8 PCR cycles. In total 384 different indexes were used. After the library preparation, the DNA concentration for 25% of the samples was measured with Qubit dsDNA BR assay. The concentrations were then averaged to determine the volume used from each sample to pool approximately 200 ng of DNA in sets of 8. To each pooled set of samples, 4 ul of the Twist mtDNA probe panel was added along with universal blockers and blocking solution, and then dried using vacuum pressure. The dried samples were then stored overnight at −20C. The following day, the capture of the mtDNA was performed manually following the Twist Target Enrichment Fast Hybridization protocol. Samples were hybridized in a thermocycler at 60C for 2 hours. For each pool of samples 15 PCR cycles were performed. DNA concentrations of the completed libraries were measured with Qubit dsDNA HS assay and size distribution was checked with the Agilent 5200 Fragment Analyzer and the High Sensitivity Large Fragment 50kb kit by the Johns Hopkins Single Cell and Transcriptomics Core. Completed libraries were pooled together at 4 nM, and sequencing was performed at 2 × 150 bp on the Illumina NovaSeq 6000 system in a single lane of a NovaSeq SP Flowcell by the Johns Hopkins Genetics Resources Core Facility High Throughput Sequencing Center. This protocol was used to sequence all follow-up visit samples. In addition, 406/1031 first visit MESA samples were sequenced to confirm the comparability of WGS and Twist sequencing. Across 183 heteroplasmies called by either WGS or Twist, the correlation of VAF was high (Pearson’s r^2^=0.91), with no excess of de novo variants seen in the Twist data relative to the WGS data (**Supplementary Figure 4**).

### Detection of mtDNA heteroplasmy

MitoHPC (20240306 version) was used for mtDNA heteroplasmy detection from WGS and Twist sequencing data^21^. MitoHPC utilizes GATK Mutect2 twice for variant identification and SAMtools for generating read count and coverage used for mtDNA-CN calculation. Prior to analyses, we downsampled each sample to 2000X at each mitochondrial genome position using the samtools *view* command. We implemented a heteroplasmy allele frequency cut-off of 5%, where variants with allele frequency 5%-95% were considered heteroplasmic, and <5% or >95% homoplasmic.

Documentation for MitoHPC can be found at: https://github.com/ArkingLab/MitoHPC. We only included variants at sites with minimal read depth in both visits was >=300 to avoid false-negative calls. We also removed INDELs and variants in the polyC homopolymer regions, and any variants with ‘base|slippage|weak|position|strand’ flag in the FILTER column of the output VCF. For multi-allelic heteroplasmy at a single position, the heteroplasmy with higher MLC score is retained. When a heteroplasmy was initially detected at only a single visit (VAF>=0.05), we determined whether a heteroplasmy was present below the detection threshold in the corresponding visit by running MitoHPC with the *freebayes*^22^ variant detector, which has higher sensitivity than Mutect2, and used the observed VAF if >0.01, or set it to a minimum VAF=0.01. We excluded a group of heteroplasmies that occurred together at similar VAF (positions 3097, 3098, and 3100), and were seen only in visit 5 in 4 MESA participants. These variants were not seen in ∼180,000 TOPMed samples nor in ∼490,000 UK Biobank samples, suggesting that they are false-positives introduced during TWIST sequencing. We also excluded position 319, which was “lost” at follow-up in 4 samples, and not seen in a Twist baseline sample despite being observed in the matching WGS data, suggesting a systematic false-negative in Twist sequencing. In the RS, we excluded a group of heteroplasmies that occurred together at similar VAF (positions 12684, 12705) that were only seen in the first visit, present in the BAM files but not detected by MitoHPC, pointing towards possible contamination.

### Sample QC

In MESA, we removed 69 samples that had DNA isolated from visit 1 using an alternate protocol than the rest of the MESA samples which was associated with significantly lower mtDNA-CN and a significant excess of heteroplasmy calls. Based on prior analyses in ∼200,000 samples from the UK Biobank^3^, we removed individuals with >5 heteroplasmies (MESA, N=3; RS, N=0). We removed 2 additional samples from MESA, 1 with homoplasmic variants mismatch between initial and follow-up visit, and 1 identified as likely contaminated by MitoHPC (Haplocheck contamination score >0.03). In RS, we remove 18 samples with haplogroup mismatches between visits. Furthermore, we excluded 1 additional sample with 5 heteroplasmic variants that were all present in the first visit but in the second visit only seen in the BAM files, pointing towards possible contamination.

### Calculation of change in heteroplasmy variant allele fraction (deltaVAF)

Given that the VAF is for a specific allele, to avoid a potential bias due to using the rCRS reference, in instances where the VAF in the first visit exceeded 0.8 or when the VAF was between 0.5 and 0.8 and a homoplasmy on that position was detected in more than 2 UK Biobank participants (based on MitoHPC alls in ∼490,000 WGS samples)^3^, we used the complement of the VAF (1 - original VAF) in both visits as the VAF in our analyses. This approach was taken as a high initial VAF likely reflects heteroplasmy of the alternate allele. Subsequently, we calculated the change in heteroplasmy variant allele fraction (deltaVAF) as: 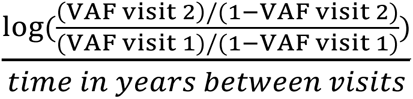.

### Association of deltaVAF with predicted variant function

We fitted generalized linear models using the R-package *glm* to determine the associations of the consequence of the mutation and the mMLC score^16^ with deltaVAF. Note that MESA had a single nonsense heteroplasmy, which was included in the missense category for analyses. Additionally, the original MLC score, which was developed solely on heteorplasmies, was modified to account for homoplasmies (mMLC), under the assumption that common homoplasmies are unlikely to be deleterious, as follows:

mMLC = MLC/(log10(homoplasmy_count+1)+1).

Homoplasmy count was derived from ∼490,000 UK Biobank participants called with MitoHPC. For analyses in MESA, we also included a covariate for sequencing batch. We performed sensitivity analyses analyzing the subset of individuals with only a single heteroplasmy, or by including only a single heteroplasmy per participant selected based on the highest mMLC score. Meta-analyses were performed using fixed-effects inverse variance weighting as implemented in the R-package *metafor*.

### Association of deltaVAF with mortality

We performed Cox Proportional Hazards models using the R *survival* package to determine the association between deltaVAF and all-cause mortality, with follow-up time starting from the time of second DNA collection. A single heteroplasmy, selected based on the largest deltaVAF, was chosen per individual to minimize potential confounding due to non-driver mutations. Analyses were adjusted for age using a natural spline with 4 degrees of freedom, sex, race, and smoking status (current, previous, never). DeltaVAF was multiplied by a factor of 10 so that hazard ratios are reported per 0.10 change in deltaVAF. Analyses for MESA also included DNA collection center and sequencing batch. Analyses in RS were truncated ten years after time of second DNA collection to enhance comparability to MESA. Secondary analyses also included mMSS calculated at the second visit from variants with VAF >=5% to assess whether the association of deltaVAF with mortality provides additional prognostic information beyond the overall heteroplasmic burden. We performed sensitivity analyses in which we used the rank-transformed deltaVAF, demonstrating that results were not driven by extreme values (**Supplementary Table 3**). Meta-analyses were performed as described above. All *P* values were calculated using a two-sided test.

### Statistics and reproducibility

Participant selection from MESA and RS are described in the Study populations section above. No statistical method was used to predetermine the sample size.

### Data availability

#### Multi-Ethnic Study of Atherosclerosis

MESA WGS sequencing data is available through dbGaP accession number phs001416.v3.p1. Twist mtDNA sequencing data has been deposited with the parent study, and is available upon request from the study coordinating center (https://www.mesa-nhlbi.org/). Due to restrictions based on privacy regulations and informed consent of the participants, data cannot be made freely available in a public repository.

#### Rotterdam Study

Rotterdam Study data can be obtained upon request. Requests should be directed towards the management team of the Rotterdam Study, which has a protocol for approving data requests. Due to restrictions based on privacy regulations and informed consent of the participants, data cannot be made freely available in a public repository.

### Code availability

Mitochondrial genetic variation was called using the MitoHPC pipeline: https://github.com/ArkingLab/MitoHPC Genetic ancestry was called in the Rotterdam Study using ADMIXTURE: https://dalexander.github.io/admixture/index.html

## Acknowledgements

This work is supported by the National Heart, Lung and Blood Institute (NHLBI) (R01HL1445609, R01HL131573) and National Institute on Aging (P30AG021334).

Whole genome sequencing (WGS) for the Trans-Omics in Precision Medicine (TOPMed) program was supported by the National Heart, Lung and Blood Institute (NHLBI). WGS for “NHLBI TOPMed: Multi-Ethnic Study of Atherosclerosis (MESA)” (phs001416.v3.p1) was performed at the Broad Institute of MIT and Harvard (3U54HG003067-13S1). Centralized read mapping and genotype calling, along with variant quality metrics and filtering were provided by the TOPMed Informatics Research Center (3R01HL-117626-02S1). Phenotype harmonization, data management, sample-identity QC, and general study coordination, were provided by the TOPMed Data Coordinating Center (3R01HL-120393-02S1), and TOPMed MESA Multi-Omics (HHSN2682015000031/HSN26800004). The MESA projects are conducted and supported by the National Heart, Lung, and Blood Institute (NHLBI) in collaboration with MESA investigators. Support for the Multi-Ethnic Study of Atherosclerosis (MESA) projects are conducted and supported by the National Heart, Lung, and Blood Institute (NHLBI) in collaboration with MESA investigators. Support for MESA is provided by contracts 75N92020D00001, HHSN268201500003I, N01-HC-95159, 75N92020D00005, N01-HC-95160, 75N92020D00002, N01-HC-95161, 75N92020D00003, N01-HC-95162, 75N92020D00006, N01-HC-95163, 75N92020D00004, N01-HC-95164, 75N92020D00007, N01-HC-95165, N01-HC-95166, N01-HC-95167, N01-HC-95168, N01-HC-95169, UL1-TR-000040, UL1-TR-001079, UL1-TR-001420, UL1TR001881, DK063491, and R01HL105756. The authors thank the other investigators, the staff, and the participants of the MESA study for their valuable contributions. A full list of participating MESA investigators and institutes can be found at http://www.mesa-nhlbi.org.

The Rotterdam Study is funded by Erasmus Medical Center and Erasmus University, Rotterdam, Netherlands Organization for the Health Research and Development (ZonMw), the Research Institute for Diseases in the Elderly (RIDE), the Ministry of Education, Culture and Science, the Ministry for Health, Welfare and Sports, the European Commission (DG XII), and the Municipality of Rotterdam. The current study was supported by VOILA (ZonMW 457001001). The authors are grateful to the study participants, the staff from the Rotterdam Study and the participating general practitioners and pharmacists.

---

**Supplementary Table 1.** Study population characteristics stratified by number of heteroplasmies.

|  | Multi-Ethnic Study of Arteriosclerosis |  |  | Rotterdam Study |  |  |
| --- | --- | --- | --- | --- | --- | --- |
|  | 0<br>(N=643) | 1+<br>(N=388) | Overall<br>(N=1031) | 0<br>(N=221) | 1+<br>(N=152) | Overall<br>(N=373) |
| **Age**<br>**(Time 1)** |  |  |  |  |  |  |
| Mean<br>(SD) | 58.7<br>(9.0) | 60.1<br>(8.8) | 59.2 (8.9) | 64.5<br>(5.54) | 66.7<br>(6.16) | 65.4<br>(5.89) |
| Median<br>[Min, Max] | 58.0<br>[44.0,<br>83.0] | 59.0<br>[45.0,<br>82.0] | 58.0<br>[44.0,<br>83.0] | 64.3<br>[55.4,<br>79.9] | 65.6<br>[55.4,<br>95.1] | 64.8<br>[55.4,<br>95.1] |
| **Sex** |  |  |  |  |  |  |
| Women | 350<br>(54.4%) | 204<br>(52.6%) | 554<br>(53.7%) | 123<br>(55.7%) | 94<br>(61.8%) | 217<br>(58.2%) |
| Men | 293<br>(45.6%) | 184<br>(47.4%) | 477<br>(46.3%) | 98<br>(44.3%) | 58<br>(38.2%) | 156<br>(41.8%) |
| **Race** |  |  |  |  |  |  |
| White /<br>European | 248<br>(38.6%) | 176<br>(45.4%) | 424<br>(41.1%) | 217<br>(98.2%) | 150<br>(98.7%) | 367<br>(98.4%) |
| Chinese /<br>Asian | 19<br>(3.0%) | 9 (2.3%) | 28<br>(2.7%) | 2 (0.9%) | 2 (1.3%) | 4 (1.1%) |
| Hispanic | 229<br>(35.6%) | 118<br>(30.4%) | 347<br>(33.7%) |  |  |  |
| Black /<br>African | 147<br>(22.9%) | 85<br>(21.9%) | 232<br>(22.5%) | 2 (0.9%) | 0 (0%) | 2 (0.5%) |
| **Smoking**<br>**(Time 1)** |  |  |  |  |  |  |
| Current | 79<br>(12.3%) | 62<br>(16.0%) | 141<br>(13.7%) | 47<br>(21.3%) | 33<br>(21.7%) | 80<br>(21.4%) |
| Never | 313<br>(48.7%) | 175<br>(45.1%) | 488<br>(47.3%) | 63<br>(28.5%) | 47<br>(30.9%) | 110<br>(29.5%) |
| Previous | 250<br>(38.9%) | 151<br>(38.9%) | 401<br>(38.9%) | 107<br>(48.4%) | 71<br>(46.7%) | 178<br>(47.7%) |
| Missing | 1 (0.2%) |  | 1 (0.1%) | 4 (1.8%) | 1 (0.7%) | 5 (1.3%) |
| **Smoking**<br>**(Time 2)** |  |  |  |  |  |  |
| Current | 53<br>(8.2%) | 39<br>(10.1%) | 92<br>(8.9%) | 48<br>(21.7%) | 28<br>(18.4%) | 76<br>(20.4%) |
| Never | 268<br>(41.7%) | 152<br>(39.2%) | 420<br>(40.7%) | 64<br>(29.0%) | 48<br>(31.6%) | 112<br>(30.0%) |
| Previous | 316<br>(49.1%) | 196<br>(50.5%) | 512<br>(49.7%) | 109<br>(49.3%) | 76<br>(50.0%) | 185<br>(49.6%) |
| Missing | 6 (0.9%) | 1 (0.3%) | 7 (0.7%) |  |  |  |
| **Time**<br>**Between**<br>**Visits**<br>**(yrs)** |  |  |  |  |  |  |
| Mean<br>(SD) | 9.39<br>(0.40) | 9.39<br>(0.37) | 9.39<br>(0.39) | 6.45<br>(0.327) | 6.43<br>(0.324) | 6.44<br>(0.325) |
| Median<br>[Min, Max] | 9.42<br>[8.10, 10.60] | 9.41<br>[7.96, 10.50] | 9.42<br>[7.96, 10.60] | 6.42<br>[5.29, 8.32] | 6.40<br>[5.17, 7.80] | 6.41<br>[5.17, 8.32] |
| **Status** |  |  |  |  |  |  |
| Alive | 579<br>(90.0%) | 335<br>(86.3%) | 914<br>(88.7%) | 146<br>(66.1%) | 101<br>(66.4%) | 247<br>(66.2%) |
| Dead | 63<br>(9.8%) | 53<br>(13.7%) | 116<br>(11.3%) | 75<br>(33.9%) | 51<br>(33.6%) | 126<br>(33.8%) |
| Missing | 1 (0.2%) |  | 1 (0.1%) |  |  |  |

| Time to Event (yrs) |  |  |  |  |  |  |
| --- | --- | --- | --- | --- | --- | --- |
| Mean (SD) | 7.75 (1.30) | 7.65 (1.45) | 7.71 (1.36) | 8.57 (2.55) | 8.58 (2.60) | 8.57 (2.57) |
| Median [Min, Max] | 8.07 [0.972, 8.68] | 8.07 [1.08, 8.68] | 8.07 [0.972, 8.68] | 10.0 [0.110, 10.0] | 10.0 [0.244, 10.0] | 10.0 [0.110, 10.0] |
| Missing | 1 (0.2%) | 2 (0.5%) | 3 (0.3%) |  |  |  |

**Supplementary Table 2.** deltaVAF stratified by extant vs. de novo.

|  | **Multi-Ethnic Study of Atherosclerosis** |  | **Rotterdam Study** |  |
| --- | --- | --- | --- | --- |
| **deltaVAF** | **Extant (385)** | **De novo (126)** | **Extant (172)** | **De novo (39)** |
| -0.10 | 22 | 0 | 5 | 0 |
| -0.05 | 38 | 0 | 14 | 0 |
| Neutral | 305 | 20 | 126 | 7 |
| +0.05 | 17 | 27 | 12 | 7 |
| +0.10 | 12 | 48 | 13 | 19 |
| +0.20 | 0 | 15 | 2 | 6 |

*Note:* Negative values are less or equal positive values are greater or equal

**Supplementary Table 3.**
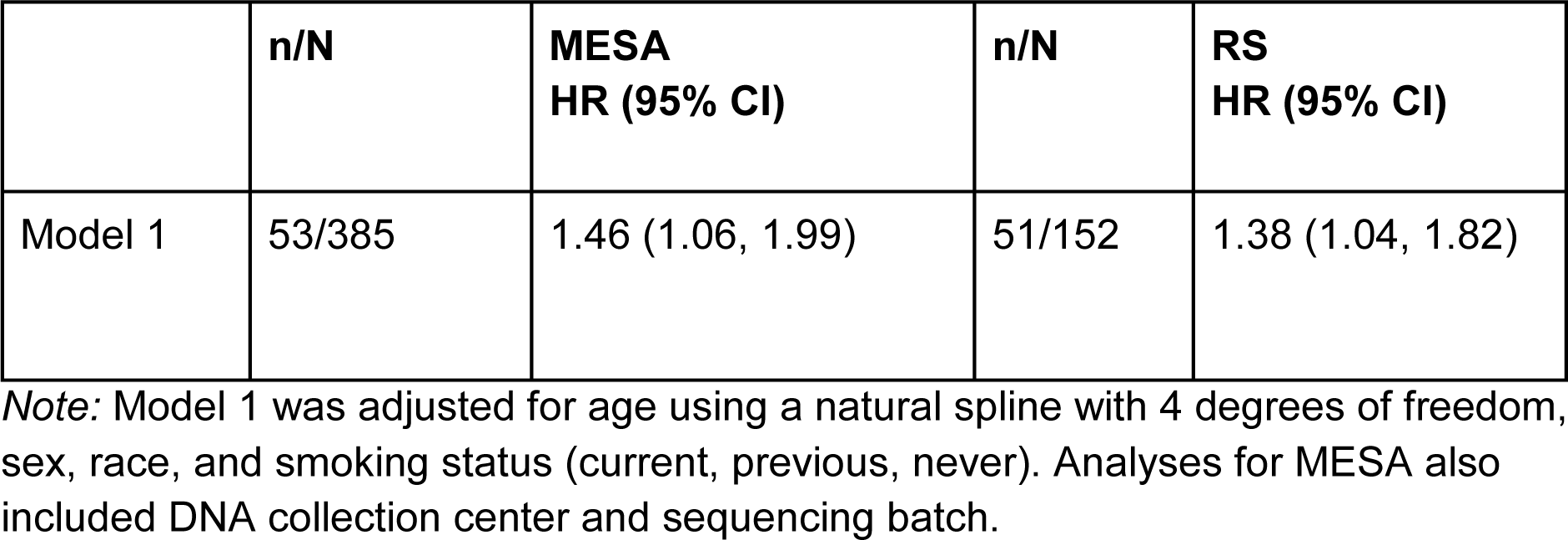
Associations between rank-transformed deltaVAF and all-cause mortality.

|  | n/N | MESA<br>HR (95% CI) | n/N | RS<br>HR (95% CI) |
| --- | --- | --- | --- | --- |
| Model 1 | 53/385 | 1.46 (1.06, 1.99) | 51/152 | 1.38 (1.04, 1.82) |

*Note:* Model 1 was adjusted for age using a natural spline with 4 degrees of freedom, sex, race, and smoking status (current, previous, never). Analyses for MESA also included DNA collection center and sequencing batch.

**Supplementary Fig 1.**
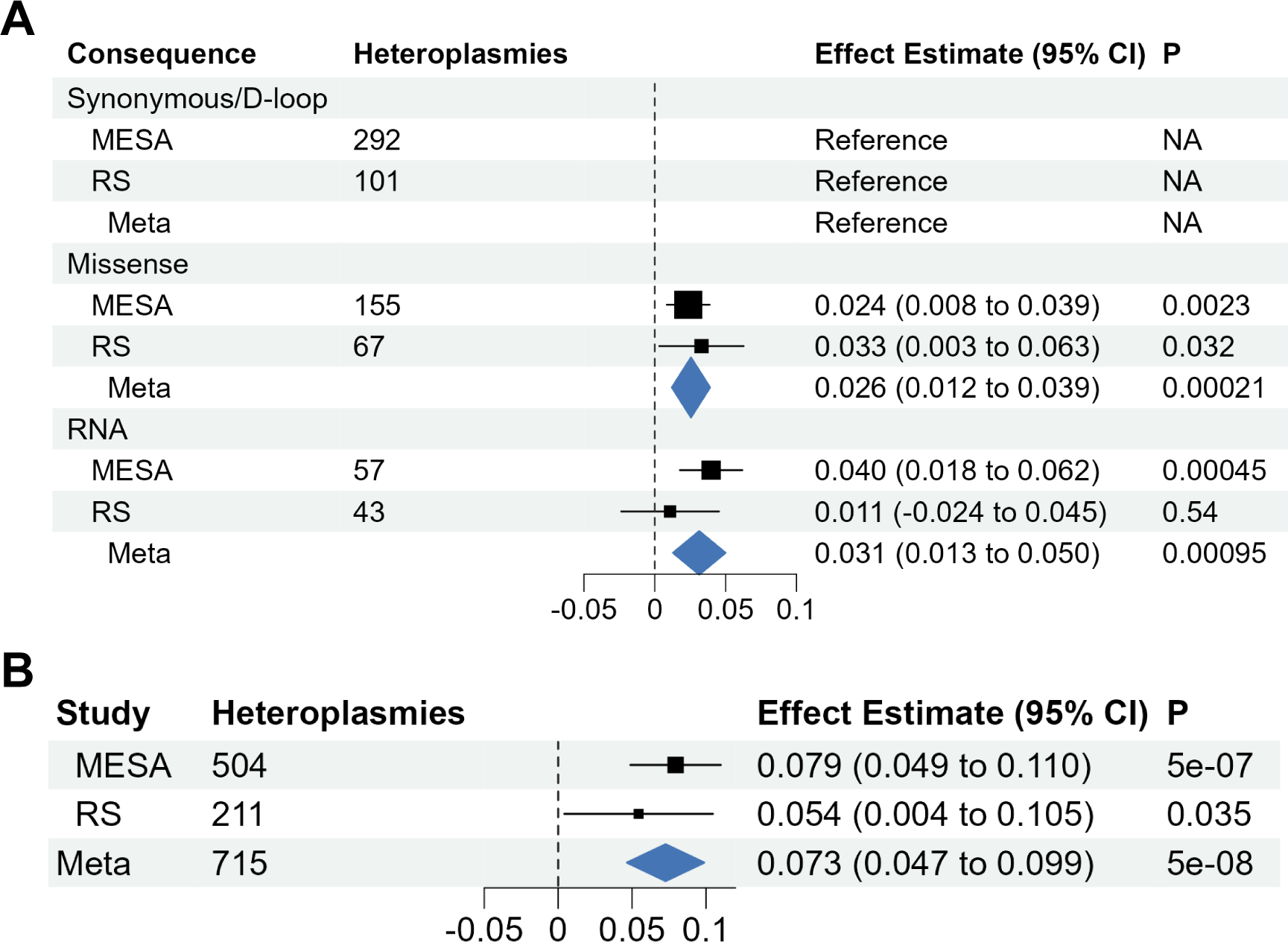
Meta-analyses of association between functional categories and functional consequence and deltaVAF. **a**, Meta-analyses of the associations of functional categories on deltaVAF. **b**, Meta-analysis of mMLC on deltaVAF

**Supplementary Fig 2.**
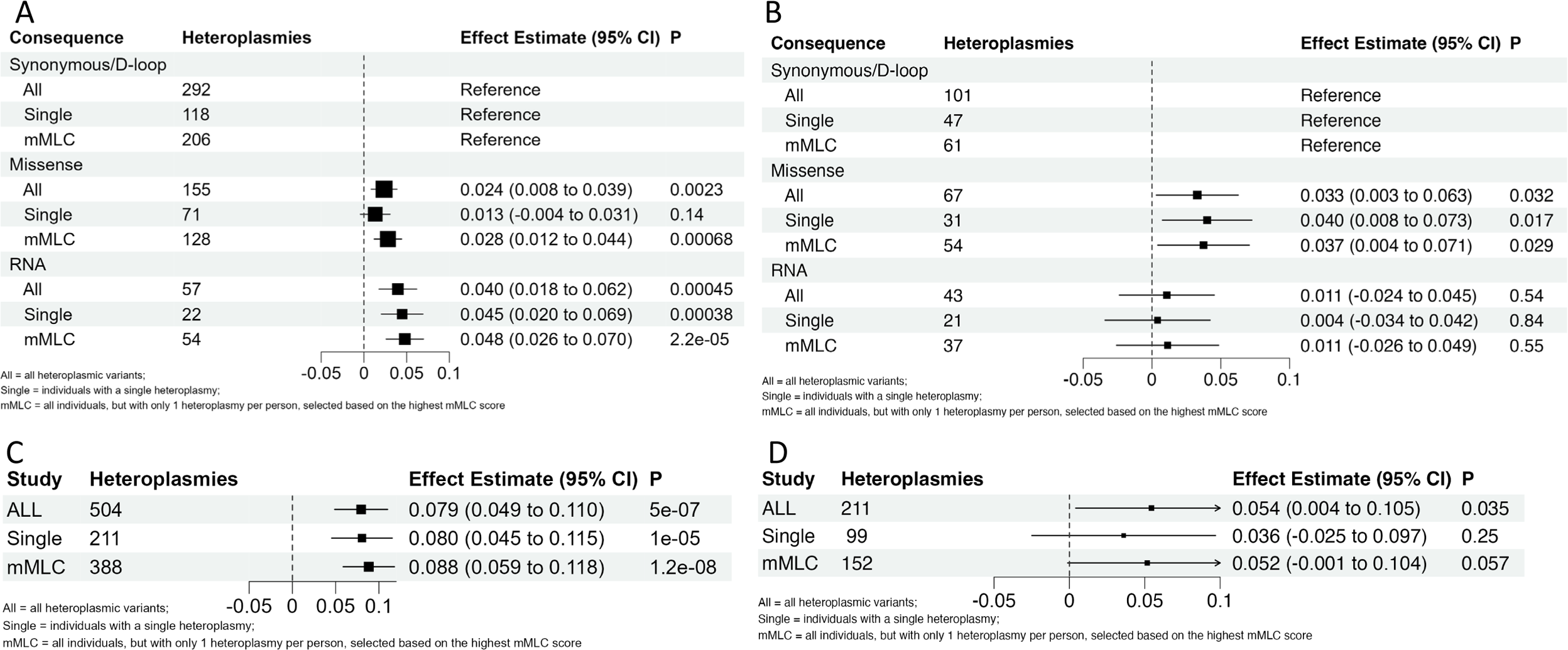
Association between functional categories and functional consequence and deltaVAF in sub-sampled cohorts. **a,b**, Associations of functional categories with deltaVAF in (**a**) MESA and (**b**) RS. c,d, Association of mMLC and deltaVAF in (**c**) MESA and (**d**) RS. ALL indicates the full cohort, Single indicates individuals with only a single heteroplasmy, and mMLC indicates all individuals, but with only 1 heteroplamsy chosen per person, selected based on the highest mMLC score.

**Supplementary Fig 3.**
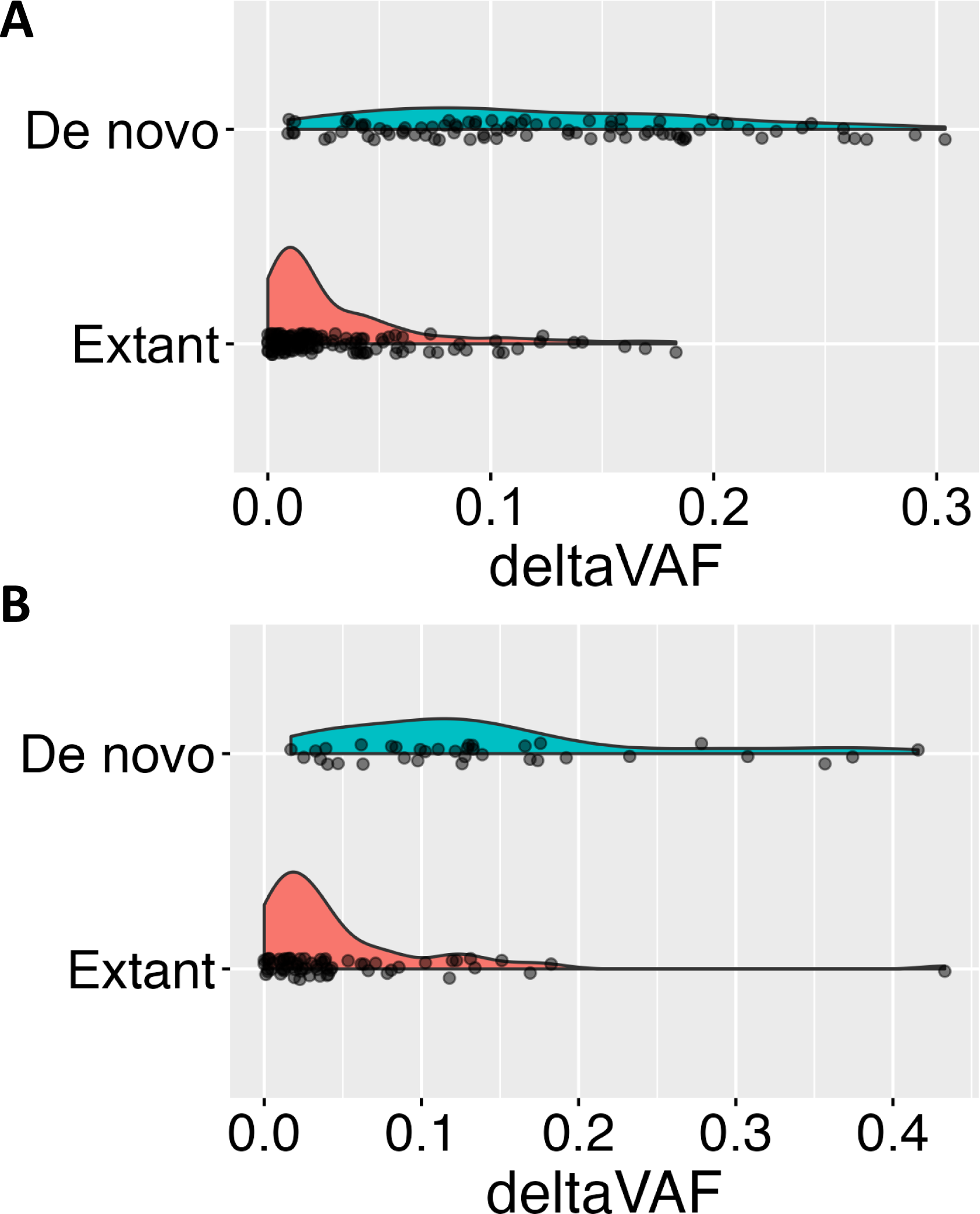
deltaVAF distribution stratified by de novo status. **a,b**, Density plots of deltaVAF stratified by de novo status in in (**a**) MESA and (**b**) RS. De novo is defined is being VAF<0.05 at baseline, and greater than 0.05 in the second visit.

**Supplementary Fig 4.**
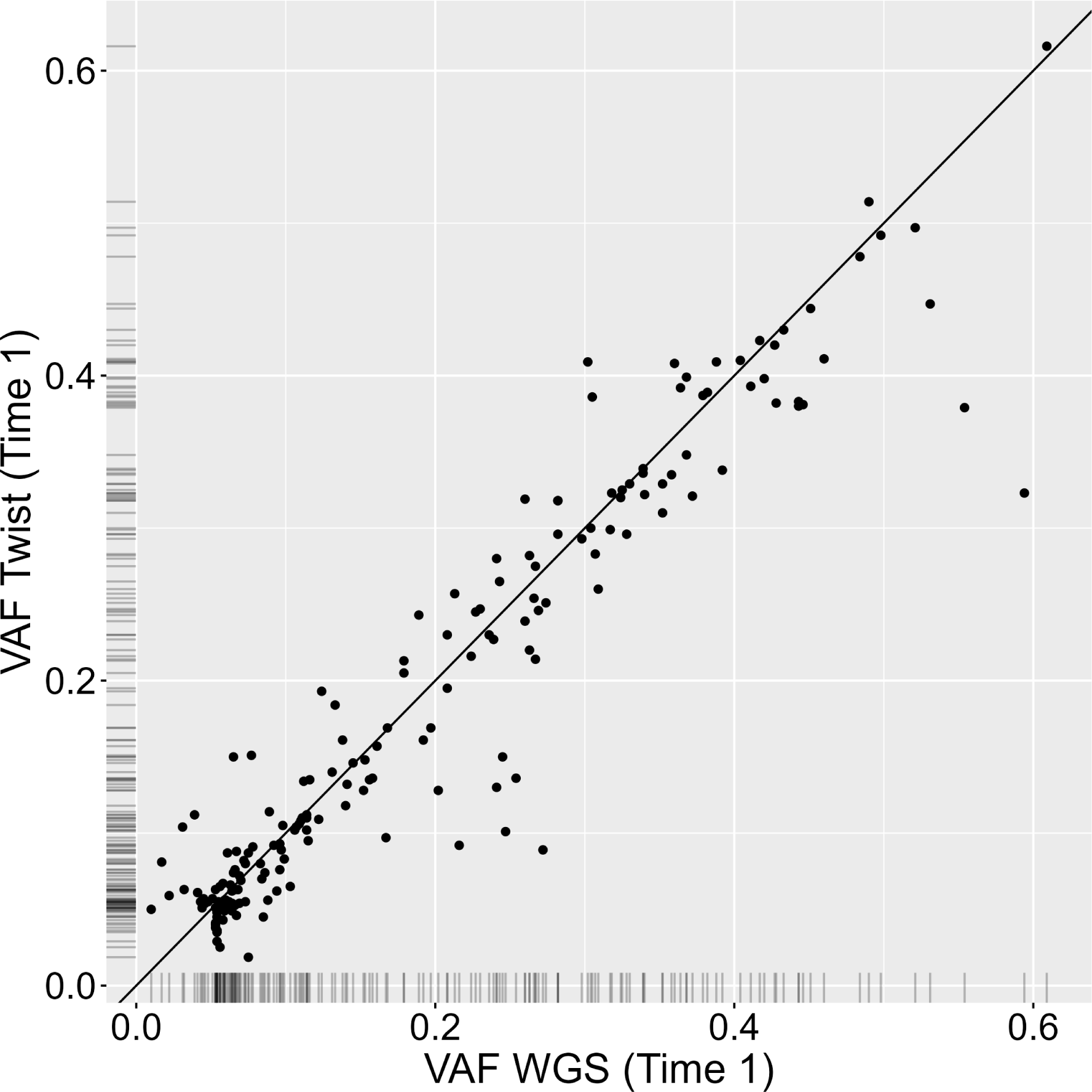
Comparison of VAF between WGS and Twist sequencing. Correlation between VAF at the first measurement (x-axis) and the second measurement (y-axis), with the black line indicating perfect correlation. Data is generated from 406 MESA samples sequenced by both technologies at the baseline visit.

